# Low-Cost, Label-Free Blue Light Cystoscopy through Digital Staining of White Light Cystoscopy Videos

**DOI:** 10.1101/2024.03.21.24304656

**Authors:** Shuang Chang, Greyson A Wintergerst, Camella Carlson, Haoli Yin, Kristen R. Scarpato, Amy N. Luckenbaugh, Sam S. Chang, Soheil Kolouri, Audrey K. Bowden

## Abstract

Bladder cancer is 10^th^ most common malignancy and carries the highest treatment cost among all cancers. The high cost of bladder cancer treatment stems from its high recurrence rate, which necessitates frequent surveillance. White light cystoscopy (WLC), the standard of care surveillance tool to examine the bladder for lesions, has limited sensitivity for early-stage bladder cancer. Blue light cystoscopy (BLC) utilizes a fluorescent dye to induce contrast in cancerous regions, improving the sensitivity of detection by 43%. Nevertheless, the added cost and lengthy administration time of the dye limits the availability of BLC for surveillance. Here, we report the first demonstration of digital staining on clinical endoscopy videos collected with standard-of-care clinical equipment to convert WLC images to accurate BLC-like images. We introduce key pre-processing steps to circumvent color and brightness variations in clinical datasets needed for successful model performance; the results show excellent qualitative and quantitative agreement of the digitally stained WLC (dsWLC) images with ground truth BLC images as measured through staining accuracy analysis and color consistency assessment. In short, dsWLC can provide the fluorescent contrast needed to improve the detection sensitivity of bladder cancer, thereby increasing the accessibility of BLC contrast for bladder cancer surveillance use without the cost and time burden associated with the dye and specialized equipment.

## 1. Introduction

More than 50% of patients who are diagnosed with bladder cancer will develop recurrence.^1–3^ Hence, once diagnosed, bladder cancer patients undergo frequent surveillance indefinitely. Complete tumor detection during surveillance is critical to the patient’s survival, as failure to detect and treat cancerous lesions, especially high-grade tumors,^4–6^ can lead to high risk of muscle-invasion and mortality.

In-office surveillance allows earlier diagnosis and the use of local anesthesia, which avoids surgical complications and the cost associated with general anesthesia. The standard-of-care surveillance procedure typically involves an in-office white light cystoscopy (WLC) during urology clinic. Unfortunately, WLC has limited sensitivity for detecting small and flat lesions, resulting in residual tumors in up to 45% of cases.^7^ The limited sensitivity is due to the similarity in morphology between small, flat tumors and healthy bladder tissue, which leads to insufficient contrast and hinders sensitive examination of the bladder.

To address the sensitivity limitations of WLC, several alternative imaging tools have been introduced and integrated into clinical practice, including narrow-band imaging.^8^ The most promising tool, blue light cystoscopy (BLC),^7,9–11^ achieves a significant improvement in the sensitivity of detection over WLC by utilizing an exogenous contrasting dye (hexaminolevulinate, Cysview^®^) that selectively accumulates in cancerous tissues and fluoresces as bright red patches against a blue background under blue light illumination.^10,12^ With the added contrast, BLC successfully reduces short-term recurrence by 14-27% and increases the detection rate of high-grade tumors by 43%, compared to WLC.^7^

While BLC offers improved sensitivity, its high cost (due to the use of specialized equipment, dye and space) and the additional time required for dye administration limit its accessibility. As a result, fewer than 5% of the hospitals in the U.S. have access to in-office BLC systems.^9^ A simple, rapid, and low-cost strategy for generating BLC images during in-office cystoscopy would increase the accessibility and affordability to BLC imaging for in-office use, bringing with it the advantage of more sensitive detection of bladder tumors that can improve patient outcomes.

Recently, machine learning methods have gained increasing interest for their potential to improve the sensitivity of bladder cancer detection.^13,14^ For example, methods like CystoNet^15^ aim to circumvent the need for BLC by providing real-time bounding box overlays of suspected tumor regions during WLC, and it can achieve good sensitivity and specificity (90.9% and 98.6%, respectively). However, such WLC-based, supervised learning strategies use training datasets that were developed using manually labeled WLC images. Unfortunately, manually labeled datasets are fundamentally limited in their utility to assist with detecting the undetectable cancers that are responsible for the low sensitivity of WLC, because they only include the subset of tumors already detectable by human eyes with WLC.

To overcome the low sensitivity of WLC for bladder cancer, it is essential to visualize tumors that are not currently visible with white light imaging. Given that BLC technology is known to significantly increase the sensitivity of detection (detecting up to 40% more tumors that are initially missed by WLC),^16^ machine learning methods that leverage information from BLC data are better poised to address WLC sensitivity limitations. In 2021, Ali et al. introduced a BLC-image-based artificial intelligence diagnostic platform for predicting malignancy, invasiveness and grade in bladder tissue, where they showed the classification against malignant lesions achieved 95.77% sensitivity.^17^ While this represents the first attempt at developing a machine-learning-based algorithm for bladder cancer detection using BLC data, the proposed diagnostic platform can only be utilized in places where BLC systems are already available. Consequently, the issue of low sensitivity of bladder cancer detection remains for the 95% offices where BLC systems are not available and unlikely to be due to recent scale-backs in manufacturing of new, in-office BLC systems.^18^

To overcome the need for new capital expenditures to perform BLC imaging, we previously proposed to use machine learning to perform image-to-image (I2I) translation, a style-transfer task rather than an object detection or classification task. This approach is a form of domain adaption, where we convert an input image from the WLC domain to the BLC domain,^19^ preserving the intrinsic content (i.e., tissue structure features) while transferring the target style (i.e., the fluorescence under blue light illumination), without the need for exogenous dye or costly equipment. This conversion is akin to virtual or digital staining -- a process that has been widely demonstrated in histology to convert unstained slides, or a simple stain such as H&E, to have the appearance of more advanced, special histochemical staining, thus saving the time, cost and environmental impact associated with processing physical stains.^20–24^ Importantly, the style-transfer approach also removes the need for manual annotation of input datasets, as the input data from the target domain serves as its own ground truth.

As with all digital staining, the underlying premise of our strategy is that structural features present in the WLC image that are too minute to be easily detected with the human eye are correlated with biochemical features that lead to fluorescent staining in BLC images. For example, early-stage bladder tumors have small papillary structures or appear as a velvety patch on the bladder wall.^25^ Under a quick glance, these small tumors may be missed due to the lack of contrast in WLC or distracting inflammation in larger regions of the bladder. However, distinct texture features (such as the velvety, carpet-like mucosa) and small changes in tissue color that are not apparent to human eyes are detectable by the machine learning model.

In an initial proof-of-concept study on the feasibility of digital staining WLC data using a standard convolutional network, we showed successful style transfer from WLC to BLC.^26^ Like most digital staining implementations,^22^ however, the machine learning model used for that work (paired I2I translation) relied on the availability of near-perfectly registered source (WLC) and target (BLC) modality image pairs, which could only be acquired with a specialized research-grade system.^27^ In real clinical scenarios using commercially available cystoscopy systems (e.g., KARL STORZ Photodynamic Diagnostic system),^28^ WLC and BLC videos are collected sequentially (i.e., a manual switch is required to change from WLC mode to BLC mode). As a result, it is not practical to obtain sufficient aligned pairs from data collected with clinical systems to train a model based on paired I2I translation.

In an effort to facilitate clinical translation of the digital staining concept for bladder cancer detection, here we introduce the use of *unpaired* I2I translation^29^ to perform the digital staining task. In so doing, we demonstrate the first use of machine learning to digitally stain WLC videos captured with standard-of-care clinical equipment (a Karl Storz Blue-light cystoscopy system with Cysview®) and transform them into BLC videos that accurately depict suspicious lesions. An additional benefit of using an unpaired algorithm is that it overcomes the difficulty in collecting paired dataset from clinical cystoscopy and can enable use of a larger number of videos for more comprehensive model development.

In the following sections, we introduce the methods for collection and preparation of the clinical dataset (Section 2.1), data pre-processing (Sections 2.2 and 2.3), a description of the model (Section 2.4) and present evaluation metrics (Section 2.5). Finally, we report the on the digital staining performance in Section 3.

To our knowledge, this is the first demonstration of digital staining on cystoscopy data collected with clinical systems. Importantly, our proposed workflow fills the current gap in bladder cancer detection by enabling improved detection sensitivity through introducing fluorescent-like contrast to WLC images while simultaneously increasing the accessibility of BLC-like images by removing the time and cost requirement for conventional fluorescent visualization. It also trumpets the potential of digital staining to bring new cost-effective options for endoscopy to clinical scenarios outside of urology that can democratize access to better healthcare.

## 2. Methods

### 2.1 Data collection and data preparation

Under approval from the Vanderbilt University Medical Center Institutional Review Board, we recruited 31 patients who were scheduled for a transurethral resection of bladder tumor (TURBT) procedure with CYSVIEW at Vanderbilt University Medical Center. From each patient, we collected a few short video clips (around one minute in length). Each video clip contained both WLC and BLC images, since the standard cystoscopy procedure requires switching between the two modalities for bladder visualization. Bladder images were taken from various regions of the bladder and, given the nature of the clinical cystoscopy tools, the WLC and BLC images did not capture the exact same regions (i.e., they were unpaired). In total, we collected 45 videos (81,000 images) using the commercial KARL STORZ Blue Light Cystoscopy with Cysview® System.

To prepare the datasets, we down-sampled the video data by saving one of every three frames (with a frame rate = 30Hz) and classified each frame as WLC or BLC image.^30^ The down-sampling processing reduced data redundancy, which occurs because of the fast imaging rate of the cystoscope. Additionally, unusable images (such as those that include surgical tools in the field of view, extremely saturated, dark, or blurry images) were discarded. In total, 4491 WLC and 3965 BLC images were used as inputs to the model with an 80/20 split, resulting in 3593 and 898 WLC images and 3172 and 793 BLC images used for training and validation, respectively. No registration/pairing was necessary for the training and validation datasets, since these images were used as inputs to an unpaired I2I model. For testing, however, we manually identified 100 near-perfectly matched WLC and BLC image pairs to allow for evaluation with a ground truth. We were thus constrained to a small testing dataset, as it was difficult for clinicians to capture similar regions and orientations of tissue with the native clinical equipment. Region mismatches in WLC and BLC videos greatly limited the number of exact overlaps between WLC and BLC tissue regions, even after image registration. However, our testing dataset was specially constructed to include a variety of healthy and cancerous tissues from several patients, ensuring that our model could be tested in a manner as representative as possible. While this meant that some data from patients whose videos were used in the test set were also included in the training and validation sets, all video frames corresponding to the five seconds before and five seconds after any given testing image were removed from the training set to avoid overlapping information that may affect our evaluation assessment. The saved images were cropped to a square that circumscribe the circular cystoscopy field of view (FOV).

### 2.2 Color Normalization

Cystoscopy videos collected via clinical systems present several challenges for this work. Besides the aforementioned challenge of collecting perfectly registered datasets for the source and target image pairs to enable validation, large variations in brightness and color that exist across the patient population and across different images collected in a single video make it difficult to perform successful training on raw images extracted from clinical videos. As shown in **Figure 1(a, d)**, both WLC and BLC images exhibit color and saturation variations. Sources of such variations include the illumination settings of the cystoscopy device (which may be auto-controlled) and conditions inside the bladder (e.g., urine and debris in the bladder can cause a color shift in BLC videos).

**Figure 1.**
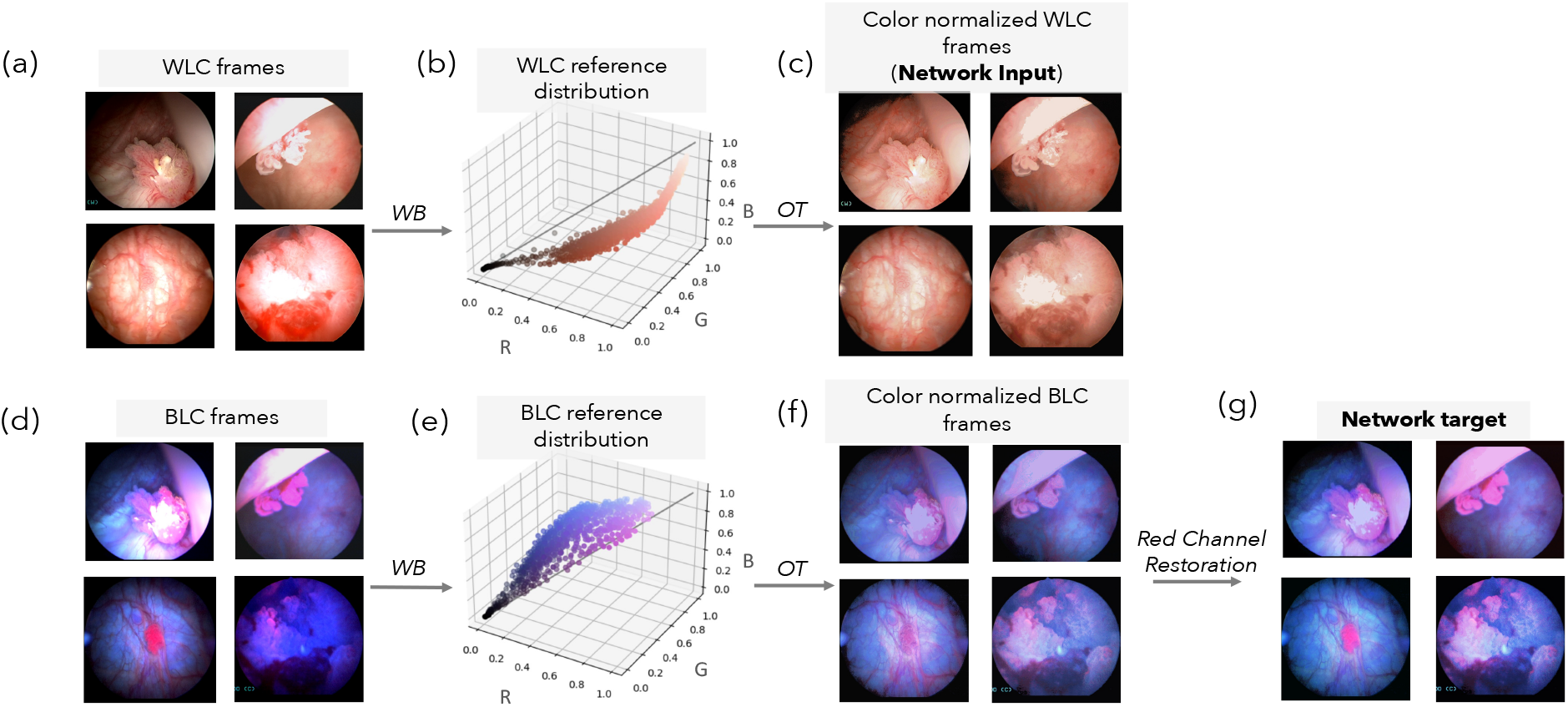
Color normalization and optimization process for WLC and BLC images to align color distributions of video datasets. A subset of the WLC/BLC images are used to find the Wasserstein barycenters (WB), which serve as the reference distributions. Original WLC and BLC images are transformed into color normalized WLC and BLC images using optimal transport. Additionally, for BLC, to restore the red channel that contain important fluorescent information, color normalized images are further adjusted to generate color-normalized BLC images with the Red Channel Restoration step, using red channel intensities in the original color channel and the relative intensity ratios between the RGB color channels.

These variable conditions can make it very difficult to appropriately predict the details needed for style transfer. Especially variable images result in out-of-distribution data, which can lead to failures of deep neural networks during both the training and testing phase. To achieve successful training on a model, in most cases, either the training set must have a homogeneous distribution or the heterogeneity of the training data must be known (e.g., using labels). While recent studies have investigated methods to generalize for out-of-distribution data,^31,32^ these primarily aim to decorrelate irrelevant features from those that are relevant. In our data, out-of-distribution cases are results of suboptimal imaging conditions, where saturation, green hues (due to urine content), and large shadowy regions mask important details. Therefore, performing normalization on every image to yield a consistent color distribution and to address the problem of saturation is a much more efficient approach, as it will yield more realistic generated images without significant imaging artifacts. In summary, our out-of-distribution data can be recovered to reveal valuable relevant features.

To account for these variations and work towards developing color-wise homogeneously distributed datasets, we first generated the color point cloud for a given image from its color distribution: each pixel was represented as a point in 3D space, where red, green and blue intensity values form the three coordinate axes, shown in **Figure 1b**. We then approached this color normalization challenge by solving an optimal transport (OT) problem:^33,34^ that is, we find found the most efficient way to transform one distribution (input image color distribution) into another distribution (a reference color distribution). Solving an OT problem requires calculation of the Wasserstein distance, which returns the minimum cost for transforming one distribution into another. We found the reference color distribution by computing the Wasserstein barycenter (WB) for WLC input distributions across several patients. The WB is defined as the distribution that minimizes its Wasserstein distance with respect to other distributions.^35–37^ In other words, the WB serves as the “reference distribution” to which we attempted to map all other images. When computing the WB, a selection of high-quality, nicely contrasted images that represent various pathological states were manually chosen to ensure that the normalized data contained high contrast and minimal artifacts that could impede visual clarity. The images came from 14 patients and were specifically selected because the images presented a wide variety of saturation, color distributions, and tissue features. With the reference distribution in hand, we then found the OT mapping between each input WLC frame and the reference distribution, transforming the input image color point clouds into color-normalized WLC datasets, shown in **Figure 1c**. Ultimately, the color-normalized WLC images would become the source data (i.e., network input) for our style-transfer process.

The same normalization process was necessary for BLC images. Hence, we similarly selected BLC images from the same 14 patients (**Figure 1d**) and created a BLC reference color distribution (Figure 2e) to produce color-normalized BLC data (**Figure 1f**). However, we found that while the color normalization step effectively unifies the brightness and color distribution for the WLC data, color-normalized BLC images showed diminished fluorescence signal, which could lead to missed tumors. This phenomenon is explainable: when optimal transport is applied on all three, color channels in the RGB color space, the red channel of each individual image is also made more uniform, contributing to the artifact. To restore the level of true tissue fluorescence associated with the red channel, we performed color normalization only on the green and blue channels of the BLC. Then we performed a Red Channel Restoration step (shown in **Figure 1g**) to compute a new red channel based on the normalized blue and green channels, wherein we first scaled the intensity distribution of the red channel in the original image by a ratiometric intensity scaling factor, which was determined by comparing the blue-channel intensities of the color-normalized and the original images. We then combined this newly scaled red channel data with the normalized green and blue channels to generate a color-normalized BLC image.

**Figure 2.**
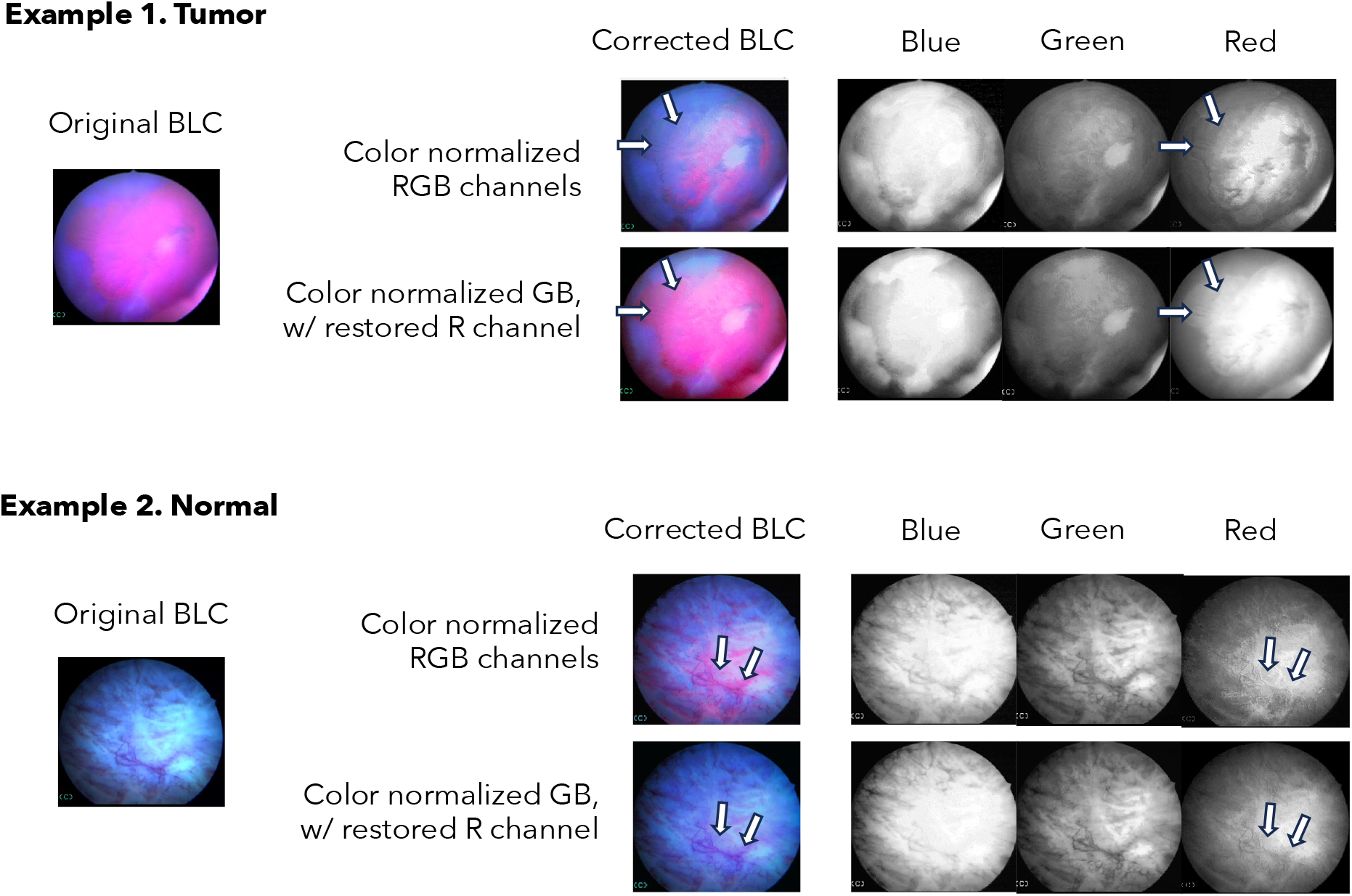
Comparison between color normalized BLC images (performed on all three color channels) vs. final color-normalized images with red channel restored illustrated in a tumor case and a normal case. Arrows show the lost fluorescence (example 1) and false fluorescence (example 2).

**Figure 2** shows two representative cases of the visual improvement gained by this Red Channel Restoration step for blue light images. In each example, the top row shows the effect of the original RGB color normalization process (as used in WLC processing), and the bottom shows the corrected image with Red Channel Restoration. In the tumor case, severe fluorescence wash-out appears in the top image, compared to the fluorescence that is restored in the post Red Channel Restoration (bottom) image. In the example of normal bladder tissue, the original color normalization led to falsely labeled fluorescence, which was removed after adjusting the red channel intensity. Arrows highlight the difference in fluorescence between the top and bottom row of each example, showing that the accurate level of fluorescence in a color-normalized image can be achieved with Red Channel Restoration step.

Following the color normalization steps described above, all WLC and BLC images used for training, validation and testing were processed to achieve color-wise homogeneous distributions.

### 2.3 Establishing a ground truth dataset for evaluation

To properly evaluate the performance of our model, we established a testing dataset that comprised WLC images for which we had identified corresponding BLC images obtained from the same location to serve as ground truth network outputs.

Because the clinically collected datasets were inherently unregistered, to generate our registered pairs, we manually identified 100 images that appeared to be near-perfectly matched by human eye. The selected image pairs were then elastically registered with MATLAB using fitgeotform2d to estimate a geometric transformation based on control point mappings between a source and target image. The control points were selected manually, where we focused on vessel patterns, tumor features and distinct bladder wall features shared by both the source image and the target image. The BLC was the source and the WLC served as the target in this case. The green channel from the WLC and the green or blue channel from the BLC (depending on the absence or presence of green-hue artifact,^30^ respectively) were used to find the transformation matrix T that enabled registration. **Figure 3** demonstrates the registration process and the associated overlays of WLC-BLC pairs before and after registration (bottom row). We calculated the mutual information (MI) index before and after the registration, between the green channels of the WLC and BLC pair, and we measured a maximum increase of 27.17 in the MI between the image pair after registration, which on average improved by 1.62 compared to unregistered WLC and BLC pairs.

**Figure 3.**
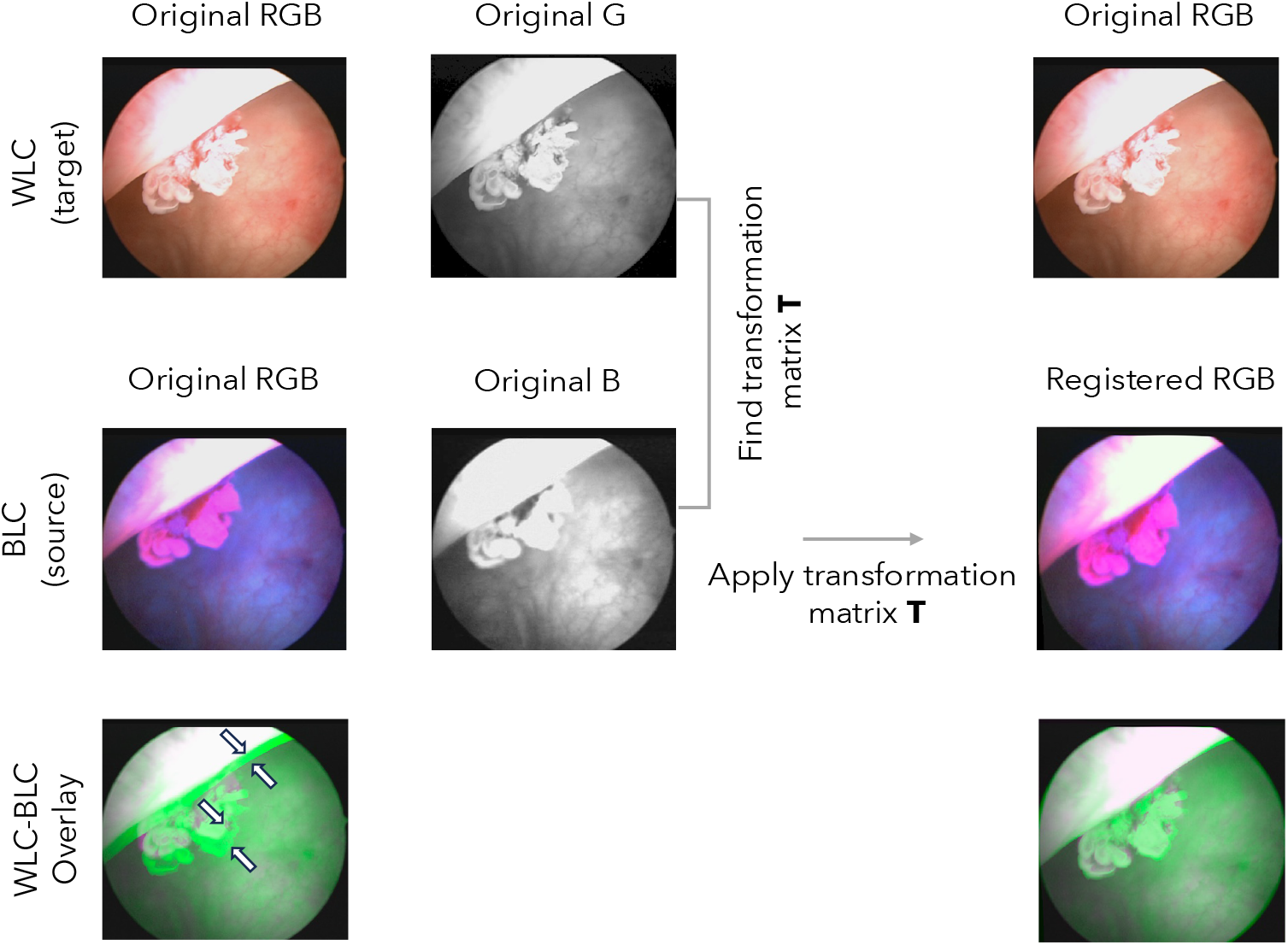
Registration of WLC and BLC pairs. Overlay images (green for WLC and purple for BLC) are shown for before (left) and after (right) registration. White arrows point to misalignment before registration. The MI improvement after registration for this image pair is 2.88.

### 2.4 Unpaired I2I model

For our unpaired image-to-image (I2I) model we used the Density Changing Regularized Unpaired Image Translation (DECENT) method, which combines a ResNet-based generator with a PatchGAN discriminator and employs autoregressive flows^38^ for density estimation (**Figure 4**).^39^ The term density here refers to the probability density of an image, which describes the areas that appear most important to the human eye through placing higher importance on edges and textures. In brief, the density-changing assumption by Xie et al. removes the restrictive one-to-one mapping assumption that is normally assumed in unpaired I2I tasks by looking at the neighboring information. The one-to-one assumption is not suitable to solve tasks where there is unequal amount of information in the two domains, as in our case. In Xie’s study, this restrictive assumption was removed by mapping the high density region of images in one domain to the high-density region of images in the other domain, and vice versa.

**Figure 4.**
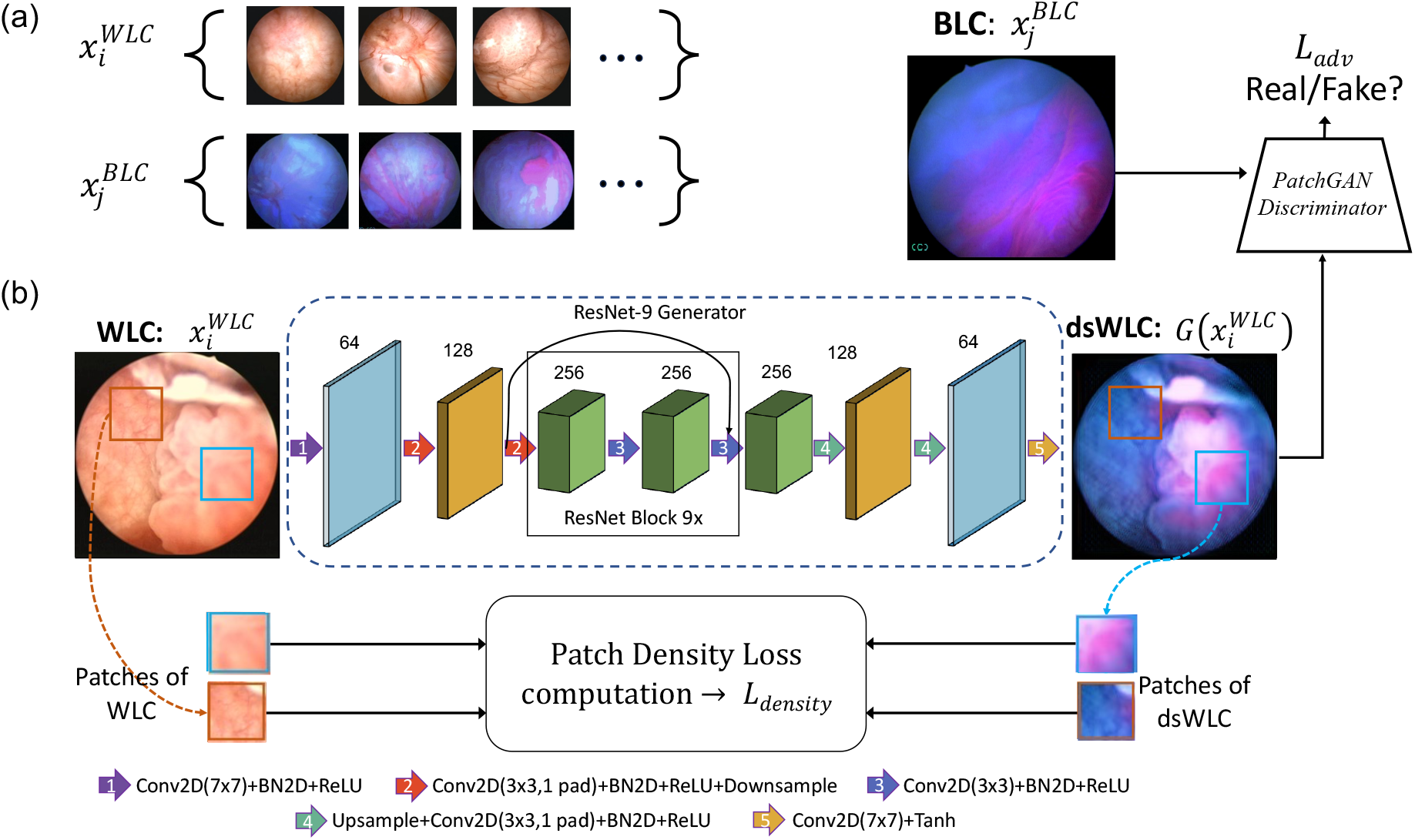
(a) Examples of unpaired input data. Each set of input data represents images that do not have corresponding, registered pairs in the other dataset. (b) The p atch-density-based GAN model utilized in our unpaired I2I. Blue dashed box represents the generator(G) which produces dsWLC images from WLC inputs.

The model consisted of three loss terms: an adversarial loss, an identity mapping loss, and a patch density loss. A least squares loss function was used for the discriminator. The loss term coefficients used in this study were the same as in the original study: 1.0, 10, and 0.01, respectively. The generator and discriminator modules were optimized using the Adam optimizer with an initial learning rate of 0.0002. The inputs to the model were color-normalized WLC 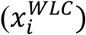 and BLC 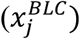 images that were resized (with a bilinear interpolation method in PyTorch) to 256 by 256 pixels from original size of 1024 by 1024, without correspondence between the WLC and BLC datasets. The outputs of the network were the digitally stained WLC (dsWLC) images, 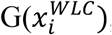, sized 256 by 256 pixels. Images were resized in this proof-of-concept study to enable direct use of the original model parameters (the original model expects a 256 by 256-sized input). In future experiments, original image sizes can be maintained to capture fine structural features to achieve better staining accuracy.

### 2.5 Evaluation metrics

To quantitatively assess the performance of the network for staining accuracy and color consistency, we defined three categories of analysis metrics. All assessments were performed by comparing the network-generated dsWLC image with the ground truth BLC image obtained from the registration process. Testing and evaluation were performed only on those image pairs for which we had BLC images as ground truth.

#### 2.5.1. Staining accuracy

Staining accuracy was defined as the percentage of correctly stained pixels of an image. To quantitatively measure staining accuracy, a fluorescence mask was developed for pixel-to-pixel evaluation. First, to remove black pixels from analysis, such as the cystoscopy viewing window surrounding each image, all pixels below a grey intensity of 40 (on a scale of 1 to 255) were excluded, resulting in image region masks such as those shown in the grey bounding box of **Figure 5**. Next, each image was analyzed in the RGB color space. Note that the fluorescent features of the tumor have a distinct red channel intensity, separate from the blue-lit background. Hence, the red to blue channel ratio (R-B) ratio was determined for each pixel in the image and used to generate a fluorescence mask. All ratios between 0.6 and 0.85 were tested in increments of 0.05, and 0.8 was empirically determined to best capture a fluorescent region using the ground truth BLC image as a reference. **Figure 5** shows representative clinically collected BLC and digitally stained images along with their corresponding image region and fluorescence masks. Because the circular field of view was distorted during the registration process, only the overlapping region between the registered BLC and dsWLC images was used in quantitative analysis.

**Figure 5.**
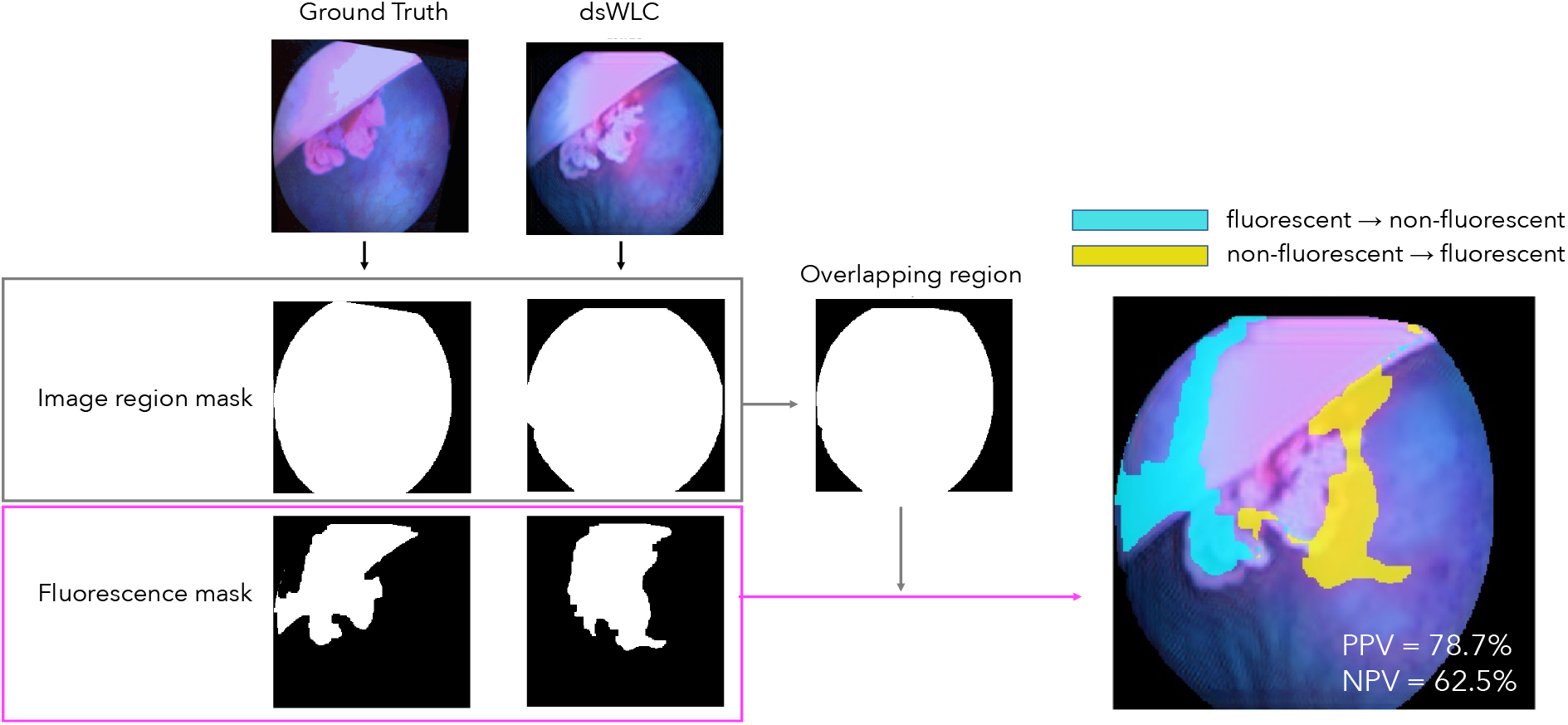
Fluorescence-segmentation-based staining accuracy assessment. Left two columns show the registered ground truth BLC, dsWLC images and their corresponding image region mask and fluorescence mask. Middle column shows the shared region between registered BLC and dsWLC. Right-most column shows overlapping-region-masked dsWLC image with overlays depending on staining accuracy: correctly stained (no overlay), incorrectly stained from fluorescent to nonfluorescent (aqua) and incorrectly stained from non-fluorescent to fluorescent (yellow). Fluorescence is determined by the R-to-B ratio threshold.

For each BLC-dsWLC registered pair, we used the mask of the BLC as the ground truth and computed the percentages of correctly and incorrectly stained pixels in the mask of the generated image. A correctly stained pixel suggests that a fluorescent (red) pixel in the ground truth also shows fluorescence in the dsWLC image, and a non-fluorescent (blue) pixel in the ground truth also appears blue in dsWLC. Incorrectly stained pixels were further divided into two cases: (1) a fluorescent pixel in the ground truth showing up as non-fluorescent pixel in dsWLC (i.e., false negative), or (2) a non-fluorescent pixel in the ground truth appearing fluorescent in dsWLC (i.e., false positive). The right column of **Figure 5** shows the incorrectly stained pixels overlaid on top of the dsWLC image, where pixels without color overlay are correctly stained pixels.

From these cases, we computed the positive predictive value (PPV) and negative predictive value (NPV) for each image, where a true positive means a fluorescent pixel in BLC appeared fluorescent in dsWLC and a true negative means a non-fluorescent pixel stayed non-fluorescent in dsWLC. Finally, we established thresholds for PPV and NPV values to declare to identify staining success for each image and evaluated the performance of digital staining on the image level for the entire testing dataset. Only tumor image pairs (i.e., the ground truth BLC image shows fluorescence) were used for the PPV calculation, as normal images had no positive regions to match.

#### 2.5.2. Color consistency

To verify the realistic appearance of the generated image, the color of the resulting dsWLC image was compared with that of the corresponding BLC image. Each pair of BLC and dsWLC images was analyzed in the RGB color space. Using the fluorescence segmentation mask, we computed the average RGB values for the fluorescent and non-fluorescent regions of each image. For BLC images, we computed average RGB values from fluorescent and non-fluorescent regions of the image using the fluorescent mask; similarly, for dsWLC images, we computed average RGB values from the union fluorescent regions and non-fluorescent regions of the BLC and dsWLC fluorescent masks, which ensures that the RGB analysis is performed on correctly stained pixels.

## 3 Results and Discussion

**Figure 6** shows examples of digitally stained WLC images from the testing data. The preliminary results demonstrated successful fluorescent labeling on tumor regions of cystoscopy images for different morphological tumor appearances. Realistic bladder wall appearance was preserved and comparable fluorescent intensities to color-normalized ground truth BLC images were generated.

**Figure 6.**
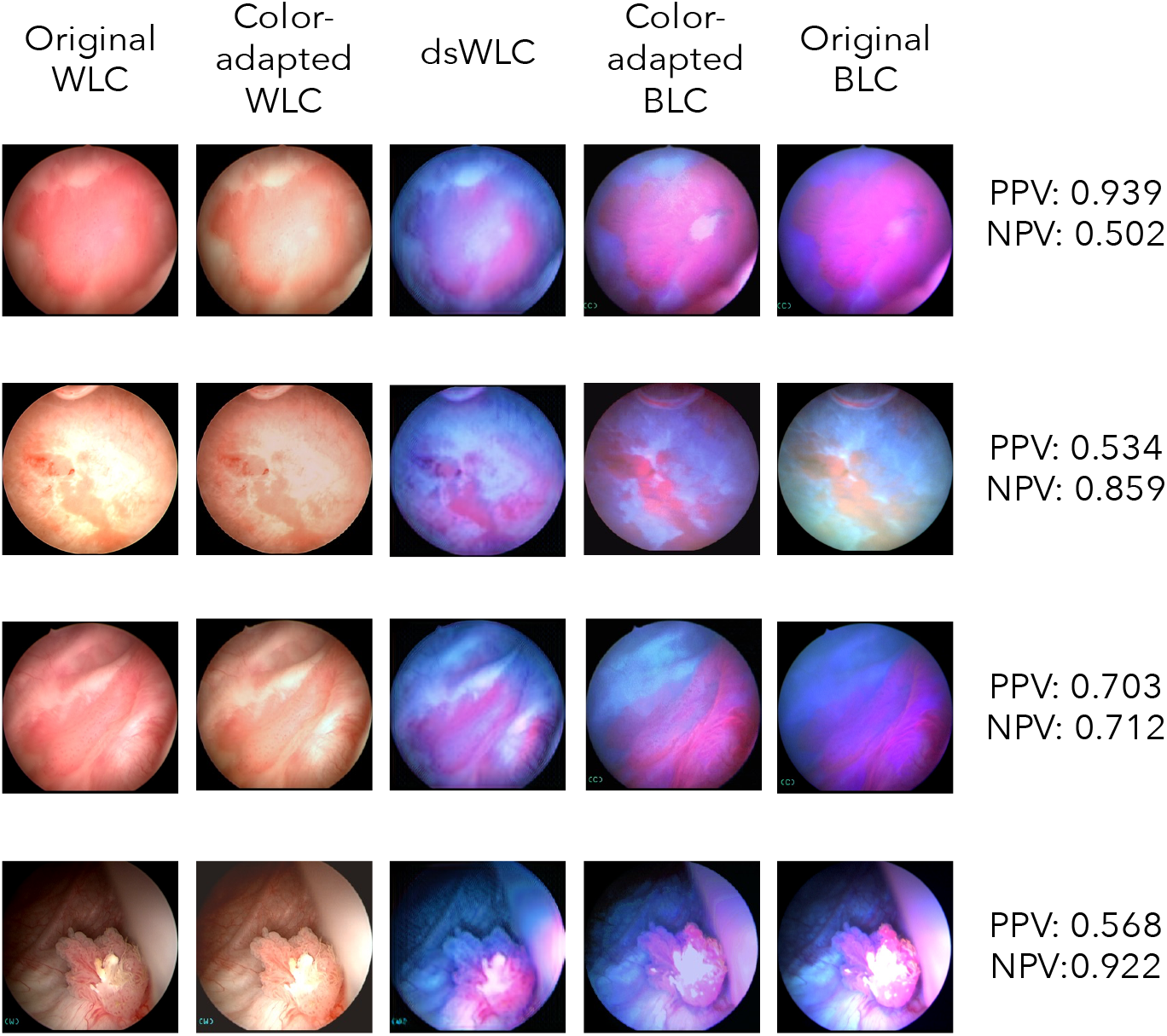
Preliminary results of digital staining in clinically collected cystoscopy data. Four examples of morphological varying bladder tumor images are presented. Digital staining was performed on color normalized WLC images (second column from the left), which produces digitally generated BLC images (middle column) that show similar appearance with the color-normalized ground truth BLC images (4th column from the left). Original WLC and BLC images are included in the left-most and right-most column, respectively.

When PPV and NPV thresholds (calculated per image) were set to 0.5, we computed 80.58% staining accuracy from the testing dataset with fluorescent masks associated with a red-blue channel intensity ratio of 0.8. We also tested other combinations of thresholds as well as various intensity ratios for fluorescent mask. The resulting staining accuracies are summarized in **Table 1**. Some inaccuracies in the digital staining can be observed in **Figure 6**. For example, false negative staining in the dsWLC images tends to occur around the outer edge of the FOV (such as row 1 and row 3). We reason that this could be due to the varying lighting conditions in the WLC image (i.e., brighter in the center, darker around the edge). Saturation in the input WLC images may lead to false fluorescence in the resulting dsWLC image (row 2). False fluorescence in the clinical, ground truth BLC is also known to be commonly found at the tangential bladder wall,^40^ and thus can lead to “false negative” regions when comparing the fluorescent masks of the dsWLC to the ground truth (row 4).

**Table 1.** Staining accuracy percentages calculated with different red to blue channel intensity ratios for the fluorescent masks.

| Red to blue<br>channel intensity<br>ratio | Staining accuracy (%) |  |  |
| --- | --- | --- | --- |
|  | PPV > 0.5 &<br>NPV > 0.5 | PPV > 0.6 &<br>NPV > 0.6 | PPV > 0.7 &<br>NPV > 0.7 |
| 0.6 | 55.34 | 47.57 | 38.83 |
| 0.65 | 64.08 | 55.34 | 46.60 |
| 0.7 | 69.90 | 65.05 | 52.43 |
| 0.75 | 78.64 | 66.99 | 61.17 |
| 0.8 | 80.58 | 74.76 | 66.02 |
| 0.85 | 74.76 | 71.84 | 67.96 |

As mentioned before, the choice of the fluorescent mask, which depends on the red-to-blue channel intensity ratio, also has an effect on the resulting staining accuracy analysis. Specifically, when the ratio was set too low, non-fluorescent regions were counted as fluorescent pixels in the mask, and thus lead to lower staining accuracy calculations. On the other hand, when the channel intensity ratio is set too high, fluorescent regions may be incompletely captured by the mask. Though higher values of the red-to-blue channel intensity ratio resulted in higher accuracy from this dataset, use of higher values may not be an accurate representation of the staining performance.

Because fluorescence contrast is critical to sensitive diagnosis of bladder tumors, we investigated the average RGB colors in fluorescence regions and normal regions of the color-normalized ground truth and generated images. The mean RGB values calculated from all images in the testing set are reported in **Figure 7**, along with the corresponding color blocks for easy visualization. As can be seen in both the color blocks as well as the RGB values, the non-fluorescent regions for ground truth and dsWLC are very similar. The fluorescent regions of the dsWLC image are slightly brighter than the ground truth, and we reasoned that this is because for dsWLC images, we computed the RGB values from the union fluorescent regions of the BLC and dsWLC fluorescent masks, and one of the main inaccuracies that occurred in the dsWLC fluorescence was at the edge of the FOV. Therefore, fewer darker fluorescent regions were captured in the dsWLC, resulting in an overall brighter average RGB measurement.

**Figure 7.**
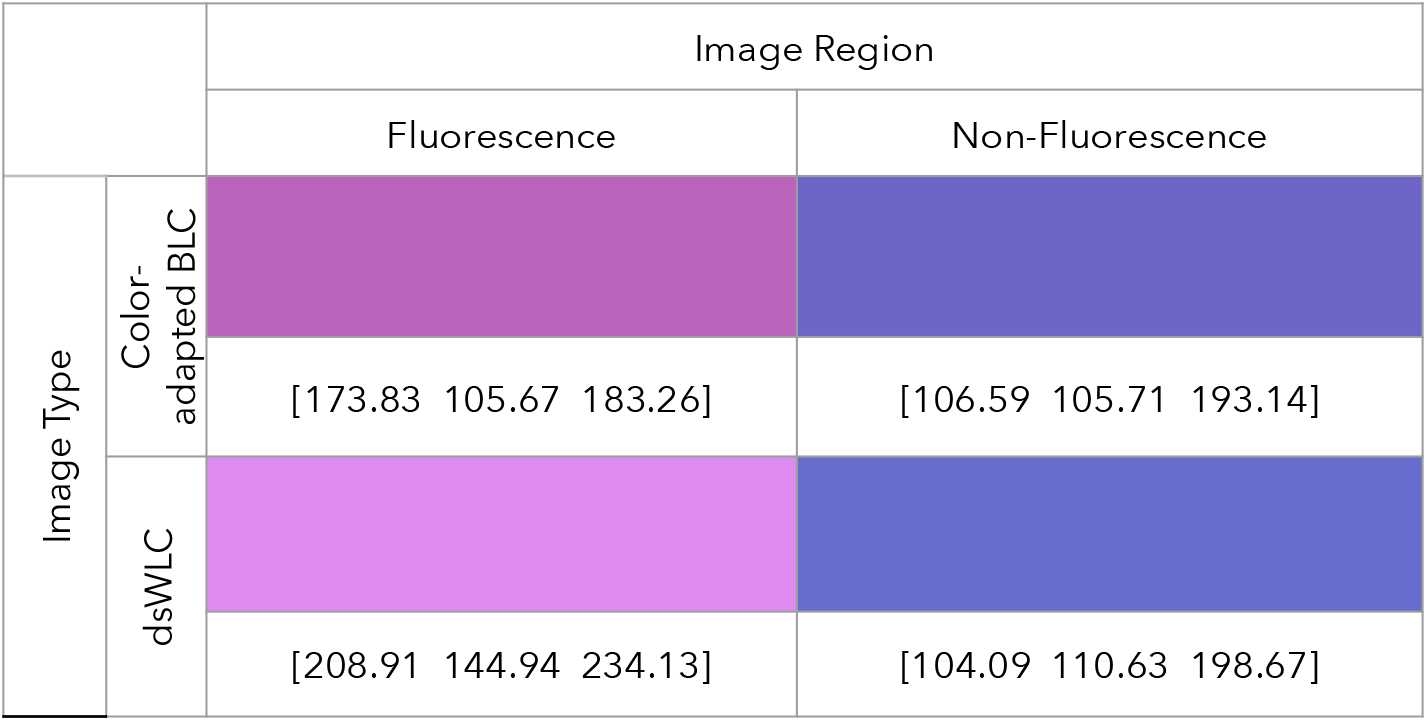
Average RGB values computed from fluorescent and non-fluorescent regions of the testing color-normalized BLC and dsWLC image pairs.

## 4 Discussion

In our current pipeline, color-normalized of the WLC is the most time-consuming step that prevents direct, real-time implementation. In the future, however, it is possible that such color modification can be built directly into the model, which will streamline the process and enable the presented work to be deployed as an accessible, cost-effective alternative to in-office BLC imaging.

While the current work aims to recreate blue light images, it is well-known that blue light imaging suffers from a high level of false positives caused by inflammation and irritations in the bladder.^40^ Here we have recreated the results of current BLC images, which means our digitally stained outputs may similarly demonstrate a high false positive rate if tested on clinical samples. We hypothesize, however, that a more comprehensive deep learning digital staining model may outperform standard BLC imaging and effectively avoid rendering benign tissues as fluorescent, since cancerous lesions have distinct textures (carpet-like or papillary appearance) and colors from the normal and benign bladder wall.^4,41^ Such features are often less distinguishable under WLC examination by human eyes, but may be sensitively and specifically detected by the model, if a large number of samples is used for training and are combined with histological confirmation results. Such a model, however, would likely require access to histological confirmation of tissue type. The current study, which aimed only to recreate BLC appearance, did not obtain histological confirmation, Instead, our assessment of the model output relied on BLC images as the assumed ground truth, which is a low-specificity imaging modality. In the future, it should be possible to improve the current model to potentially achieve digital staining with better specificity than the BLC, by incorporating additional inputs to the network and introducing additional loss terms, such as a texture map that captures the unique tissue textures that are present on tumor regions. For example, false positive staining of tangential bladder walls is a well-known aberration in conventional BLC^40^ that could be corrected for in the model, yielding an improvement in specificity over clinical BLC.

## 5 Conclusion

In summary, we successfully demonstrated digital staining of clinically acquired WLC datasets for the first time to produce dsWLC images as a quick, low-cost alternative to BLC imaging and the use of exogenous contrasting agents. The study produced successful outcomes with a 80.58% staining accuracy and minimal changes in color channel intensities to the original BLC images, suggesting the promise and feasibility of the proposed method to be implemented in a larger clinical study. The result of our study paves the way for a cost-effective alternative of BLC for in-office examinations, increasing bladder cancer detection sensitivity and removing the cost, space and time requirement for the instillation of the physical dye needed to perform BLC.

Introducing the digital staining workflow to clinical practices would increase accessibility of BLC imaging, leading to more sensitive tumor detection, thereby reducing the number of recurrences caused by failure to detect tumors under WLC imaging and lowering patient’s risk of experiencing muscle invasion and mortality. We expect that future efforts to recruit more patients and obtain histological information can lead to increased accuracy of labeling CIS tumors, contributing to solving one of the most vexing problems in bladder cancer detection.

## Data Availability

All data produced in the present study are available upon reasonable request to the authors.

## 6 Acknowledgements

We thank Ali Abbasi, Ashkan Shahbazi and Zihao Wu for helpful conversations during the initial stages of this work.

## Notes

### Competing Interest Statement

The authors have declared no competing interest.

### Funding Statement

This study was funded by Vanderbilt University faculty fund.

### Author Declarations

This study has been approved by Vanderbilt University Medical Center Institutional Review Board. IRB #211206

